# Factors Influencing COVID-19 Vaccine Acceptance and Hesitancy among Pregnant Women in Nigeria

**DOI:** 10.1101/2025.11.03.25338963

**Authors:** Sophia Osawe, Yusuff Olasunkanmi, Mobolaji Modinat Salawu, Felicia Okolo, Sikiratu Babamale, James Onyemata, Tolulope Adenekan, Adam Abdullahi, Alash’le Abimiku

## Abstract

**Background:** Vaccine hesitancy has been a serious public health concern. Pregnant women are at higher risk of developing complications related to COVID-19 infections and other infectious diseases. These complications can lead to poor outcomes for infants. This study assessed the level of hesitancy of COVID-19 vaccination among pregnant women in Nigeria.

**Methods:** A hospital-based cross-sectional study was conducted among 104 pregnant women at the Plateau State Specialist Hospital antenatal clinic. A semi-structured data collection tool of 15 items was used to assess participants’ COVID-19 vaccine hesitancy, with scores below the mean indicating hesitancy. Data were collected and managed using the Research Electronic Data Capture (REDCap). Descriptive analyses and binary logistic regression were done using Stata MP 18, and a significant p-value was set at <0.05.

**Results:** The mean age of the participants was 30.3 (SD 6.3) years, with less than half (49%) falling within the 25-34 age group. Majority (96.2%) of the participants were married, resided in the urban area (84.6%) and were employed (64.4%). More than half of the participants had more than one child (72.1%), were healthy (55.8%) and had no family members or friends who had taken the COVID-19 vaccine (70.2%). About 71% of the participants had a positive perception of the COVID-19 vaccine. The overall mean score for vaccine hesitancy among participants was 3.0 (S.D 1.6), with 57.7% hesitating to the COVID-19 vaccine. Participants with more than one child (aOR = 3.31, 95%CI=1.17-9.42), in healthy condition (aOR = 3.95, 95%CI=1.55-10.07), had family and friends who had received COVID-19 vaccine (aOR = 3.27, 95%CI=1.07-10.00) and Negative perception on COVID-19 vaccine (aOR = 4.16, 95%CI=1.05-16.57) had more likelihood to the hesitancy of COVID-19 vaccine.

**Conclusion:** Nearly three out of five pregnant women in our study expressed COVID-19 vaccine hesitancy. Pregnant women were more likely to be hesitant to COVD-19 vaccines if the felt their health was good and had a family member that had received a COVID-19 shot. Public health efforts and education campaigns for pregnant women are needed to change their perception patterns in promoting vaccination uptake and inclusion in vaccine trials.

## Introduction

The Coronavirus 2019 (COVID-19) pandemic, one of the deadliest communicable diseases in recent history, resulted in over 7 million deaths worldwide (World Health Organization, 2025a). Elderly, pregnant and lactating women, and those with underlying medical conditions are at higher risk of complications, hospitalisation, and mortality due to COVID-19. This could be attributed to possible barriers that limited access to care during the pandemic (Basoulis et al., 2023). In pregnancy, COVID-19 increases the risk of adverse birth outcomes, including preterm birth (Xu, K et al., 2024), still birth, Intensive Care Unit (ICU)admission, low birth weight (Xu, K et al., 2024) (Fleming-Dutra et al., 2023; Wei et al., 2021) and poor maternal outcomes (Kalafat et al., 2022; Wei et al., 2021).

Vaccination is a critical public health strategy to reduce the risk and spread of infectious disease and protect the health of the populace (World Health Organization, 2025b). Vaccines are important to reduce morbidity and mortality associated with infectious diseases like COVID-19, especially in high-risk populations and developing countries. High uptake of vaccines by the population increases the herd immunity threshold and thus limits the spread of infection (Haynes, 2021). COVID-19 vaccines were developed to combat the ravaging infections during the pandemic, due to the unavailability of an effective treatment options (Toubasi et al., 2022). COVID-19 vaccination during pregnancy is safe for the mother irrespective of immunosuppression status, and found to be protective against COVID in infants (Alirezaylavasani et al., 2024; Dagan et al., 2021; Fleming-Dutra et al., 2023). There is evidence of durable transplacental transfer of antibodies from COVID-19 vaccination (Kalafat et al., 2022). Beyond reducing mortality and morbidity among pregnant women, COVID-19 vaccines improve mental wellbeing and reduce the cost of hospitalization (Yussuph et al., 2023). However, despite the effectiveness of COVID-19 vaccine, there has been persistent low uptake of the during pregnancy in low/middle-income countries (LMICs), especially sub-Saharan Africa (Ashkir et al., 2023). Similarly, the condition of vaccine hesitancy has been reported among women with underlying immunosuppression like infection with HIV (Ashkir et al., 2023).

Prior to the COVID-19 pandemic, there has been documented concern about vaccine hesitancy (MacDonald & the SAGE Working Group on Vaccine Hesitancy, 2015). COVID-19 vaccine hesitancy is attributed to misinformation about the vaccines, safety of the foetus and the mother, misconceptions, mistrust of government, distrust in government and public health agencies, etc (Ashkir et al., 2023; Dirie et al., 2024; Ghamri et al., 2022; Yussuph et al., 2023). Furthermore, regional differences have been established about vaccine hesitancy in Nigeria. Northern Nigeria has constantly reported low uptake of vaccination.

Understanding the drivers of COVID-19 vaccine hesitancy among pregnant women is vital to create necessary awareness and interventions for it and other vaccine-preventable diseases. Thus, this study aims to document the social and cultural determinants of COVID-19 vaccine hesitancy among pregnant women in Jos, Northern Nigeria.

## Method

### Study setting and design

A facility-based cross-sectional study design was conducted among pregnant women who attended the antenatal clinic in Plateau State Specialist Hospital (PSSH) from January to July 2024. PSSH is located in Jos North Local Government Area of Plateau State. It has an area of 1,020 km^2^ and a population estimate of 3,206,531 (National Population Commission, 2015). It is a 300-bed tertiary health institution that specializes in medical services, trains health professionals, and serves as a referral health centre. Their antenatal clinics are on Tuesdays and Thursdays. The antenatal clinic typically begins with an interactive health talk led by a qualified community health nurse, which lasts at least 25 minutes. After the health talk, routine services, which include weight and height measurement, blood pressure estimation, urinalysis, haemoglobin estimation, and multivitamin supplementation, are carried out. Following that, pregnant women are requested to see their physician individually for a medical examination and treatment.

### Study population, sample size and sampling technique

The study population consisted of pregnant women aged 18 years and above. A total of 104 pregnant women were consented and enrolled at the antenatal clinic. Participants’ information collected includes COVID-19 vaccine hesitancy, knowledge of the COVID-19 vaccine, perception towards COVID-19, vaccination history, and socio-demographic characteristics.

### Study variables

The outcome variable for this study was hesitancy about the COVID-19 vaccine. Hesitancy on COVID-19 vaccine was measured using ten (10) semi-structured questions with a response of ‘yes’, no’, and ‘don’t know’. The ‘don’t know’ responses were merged with negative responses and scored zero (0), and the positive responses scored one (1). The scores were summed together, and a mean score of seven (7) was used in categorizing hesitancy into hesitant and non-hesitant. The explanatory variables in this study were participants’ socio-demographic which were age grouped into (≤24, 25–34, ≥35 years), sex (male and female), marital status (single and married), Parity (nulliparous (no living child) and parous (have one or more children)), current health status (not healthy and healthy), family and friend that has taken the COVID-19 vaccine (yes and no), knowledge on COVID-19 was measured using thirty-six (36) questions with a ‘yes or no responses’, each correct response was scored one, the scores were sum together and a mean score of twenty-three (23) was used in categorising the knowledge into poor and good knowledge, perception on COVID-19 vaccine was measured using a total of seventeen (17) questions with a response of ‘yes’, no’, and ‘don’t know’, the don’t know and no were merged together has zero and yes was scored one (1). The scores were summed together, and a mean score of eight (8) was used in categorizing perception into positive and negative.

### Data analysis

Data for the participants was downloaded from the REDCap database at Microsoft Excel format and then exported into Stata MP 17 for data cleaning and recoding. For analysis, ‘hesitancy’ was coded one, and ‘non-hesitancy’ was coded zero. Categorical variables were presented using frequencies and percentages, while mean and standard deviation were used to present continuous variables. The association between the outcome and explanatory variables were tested using Chi-square and Fisher’s exact test (for variables that do not meet the assumption of Chi-square). Significant variables were included in binary logistic regression to investigate the predictors of viral suppression among participants at a *p*-value < 0.05.

## Results

### Socio-demographic Characteristics

The mean age of the participants was 30.3 (6.3) years, with about 49% being between the age group of 25-34 years. More than half of the participants were employed (64.4%), lived in urban areas (84.6%), and had secondary level of education and below (55.8%). Most participants were married (96.2%), had given birth (72.1%), and belonged to households of four or fewer members (58.7%). More than two-third (89.4%) of the participants did not travel during the COVID-19 pandemic and had no family member who had received the COVID-19 vaccine (70.2%). About 63% of the participants were tested HIV-negative at the antenatal clinic. (Table 1)

**Table 1:** Frequency distribution of participants’ socio-demographic characteristics.

| **Variables** | **N (104)** | **%** |
| --- | --- | --- |
| **Age (years)** |  |  |
| ≤24 | 25 | 24 |
| 25-34 | 51 | 49 |
| ≥35 | 28 | 27 |
| Mean±SD | 30.3±6.3 |  |
| **Employment** |  |  |
| Employed | 67 | 64.4 |
| Unemployed | 37 | 35.6 |
| **Residence** |  |  |
| Urban | 88 | 84.6 |
| Rural | 16 | 15.4 |
| **Education** |  |  |
| Secondary and Below | 58 | 55.8 |
| Above secondary | 46 | 44.2 |
| **Marital status** |  |  |
| Single | 4 | 3.8 |
| Married | 100 | 96.2 |
| **Parity** |  |  |
| No child | 29 | 27.9 |
| One or more children | 75 | 72.1 |
| **Household size** |  |  |
| ≤4 | 61 | 58.7 |
| >4 | 43 | 41.3 |
| **Travelled out of the Residential area during COVID-19** |  |  |
| No | 93 | 89.4 |
| Yes | 11 | 10.6 |
| **Current Health Status** |  |  |
| Not healthy | 46 | 44.2 |
| Healthy | 58 | 55.8 |
| **Family members or friends taken the COVID-19 vaccine** |  |  |
| No | 70 | 70.2 |
| Yes | 31 | 29.8 |
| **HIV status** |  |  |
| Negative | 65 | 62.5 |
| Positive | 39 | 37.5 |

The study observed that most of the participants got information about COVID-19 from Television stations (62.5%), Radio (56.7%), Family and friends (52.9%), Health facilities (48.1%), neighbourhood (36.5%), and the Internet or social media (25%). Fig 1

**Fig 1.**
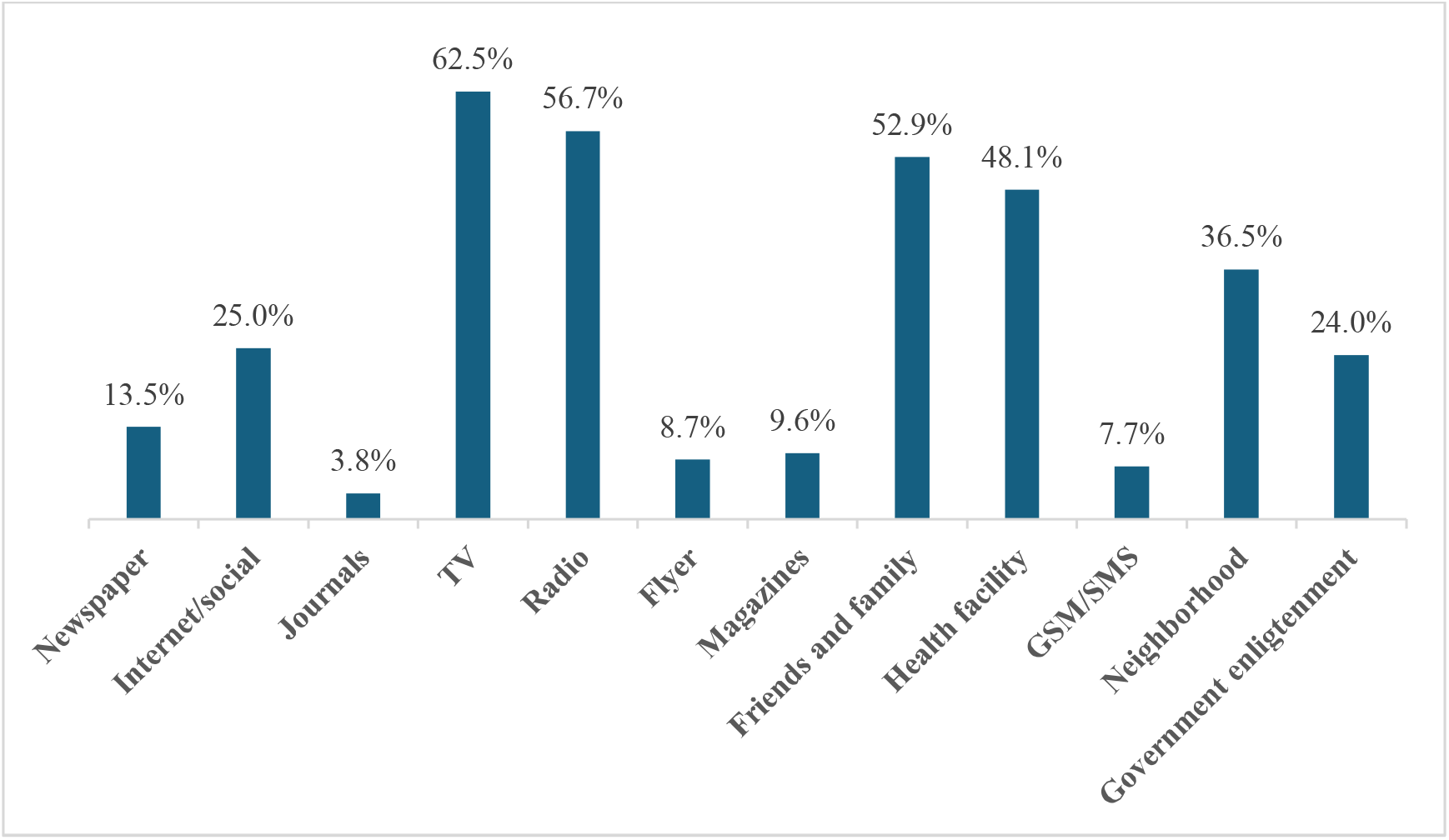
Source of Information on COVID-19.

In this study, most of the participants were aware that cough (89.4%), fever (65.4%), difficulty in breathing (55.8%), fatigue (38.5%), sore throat (34.6%), and muscle pain (24%) are symptoms of COVID-19. (Fig 2). Additionally, 74% of the participants had poor knowledge, 71.2% had a positive perception, and 57.7% were hesitant about the COVID-19 vaccine (Fig 3A, 3B, 3C).

**Fig 2.**
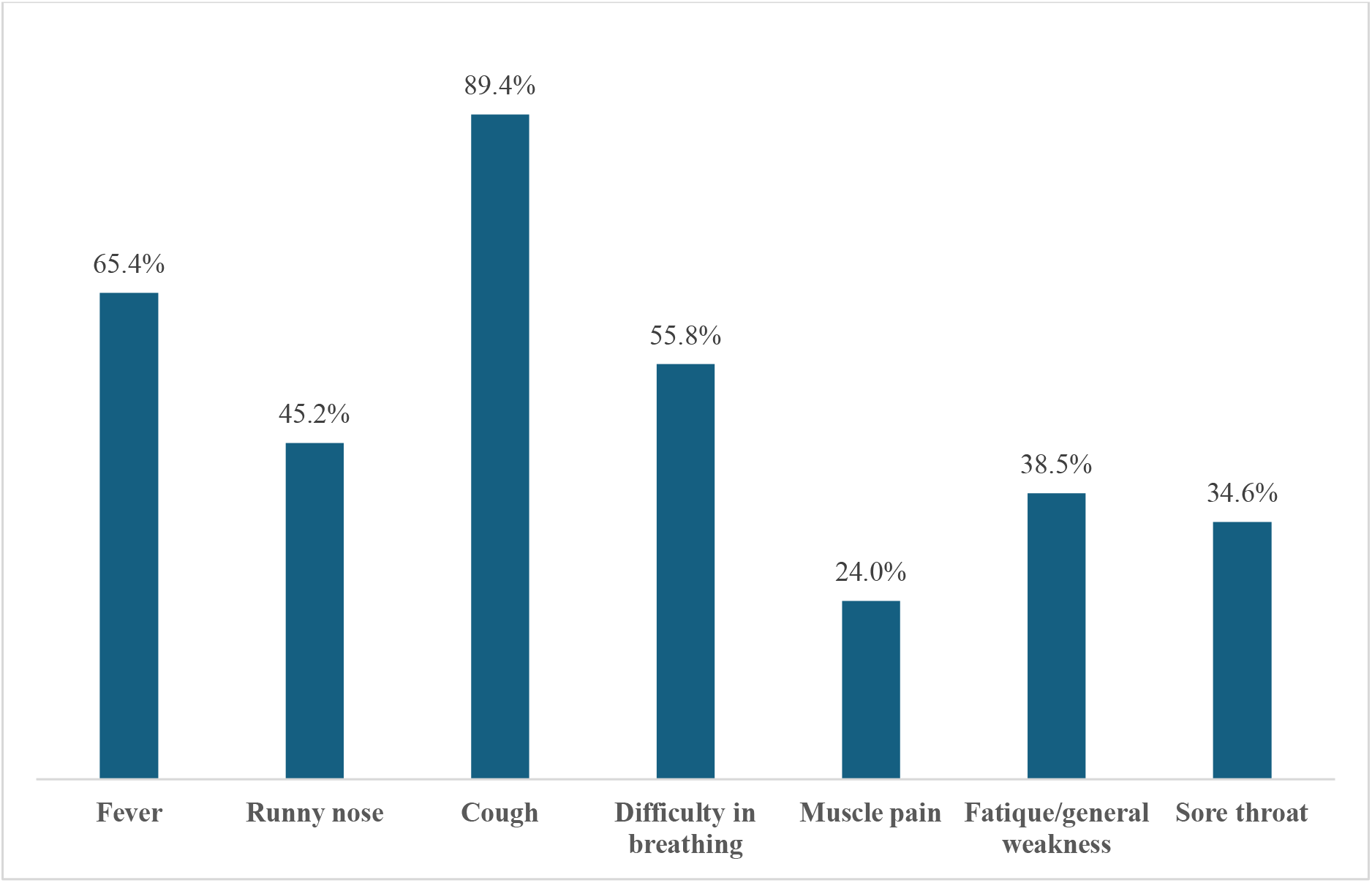
Signs and Symptoms of COVID-19.

**Fig 3.**
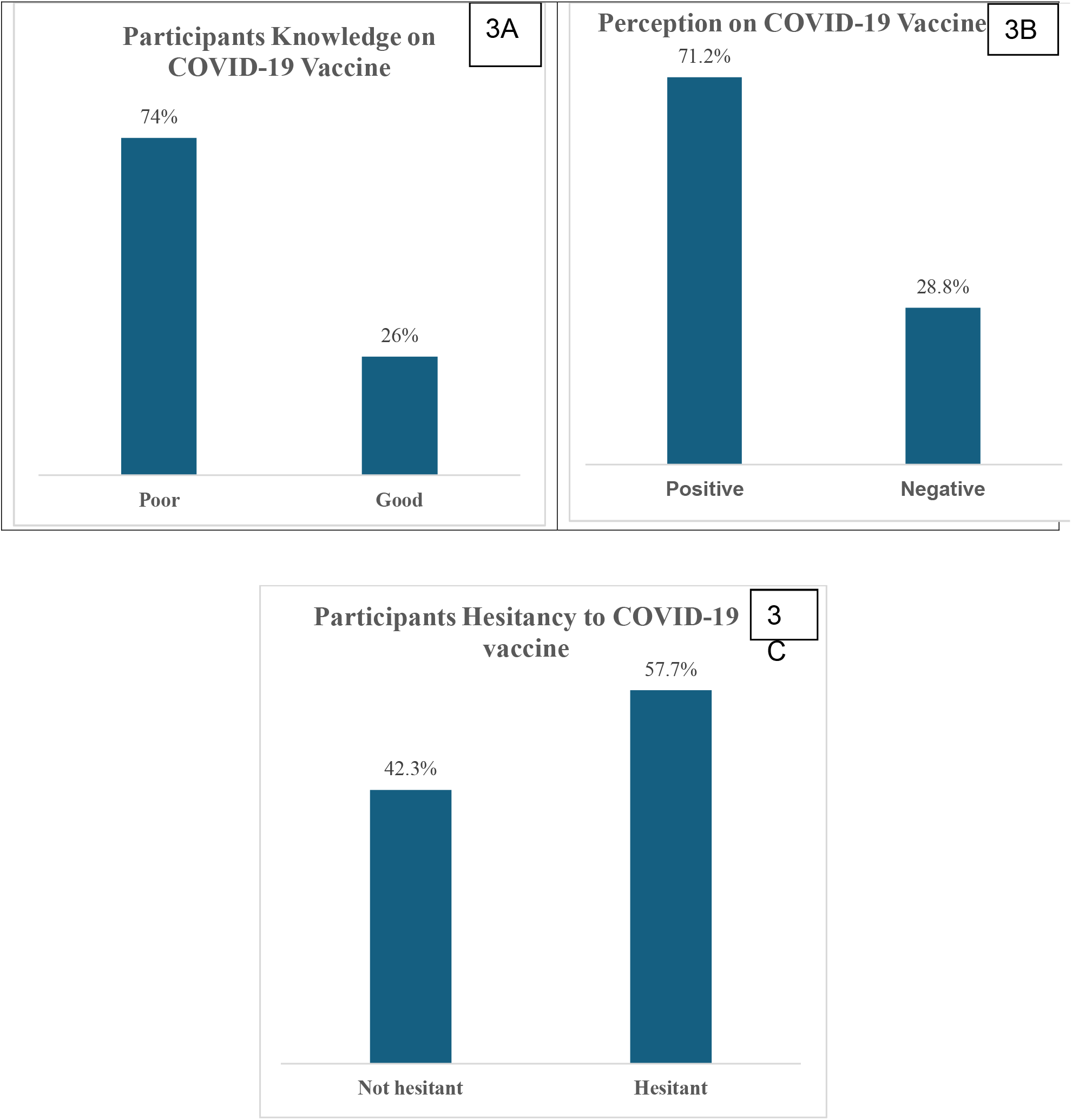

### Association between participants’ socio-demographic characteristics and COVID-19 vaccine Hesitancy

Table 2 shows variables that were significantly associated with a higher proportion of hesitancy to COVID-19 vaccine. These variables are unemployed participants (*p* = 0.165), participants that are nulliparous (*p* = 0.123), participants who did not travel during COVID-19 (*p* = 0.113), participants with good health status (*p* = 0.009), participants who had family and friends that have taken the COVID-19 vaccine (*p* = 0.086), participants with positive perception towards COVID-19 vaccine (*p* = 0.016), participants with good knowledge on COVID-19 vaccine (*p* = 0.044).

**Table 2:**
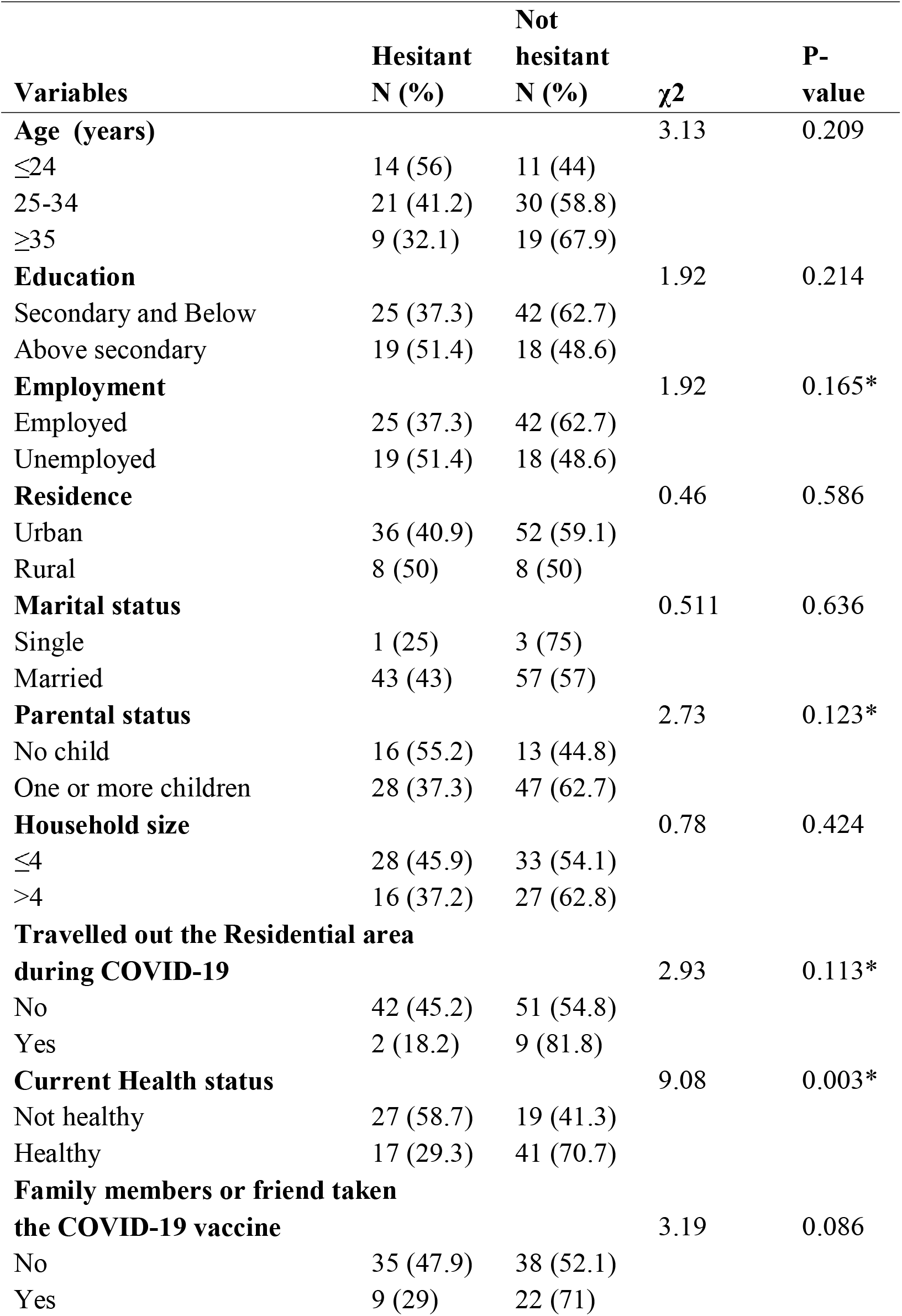
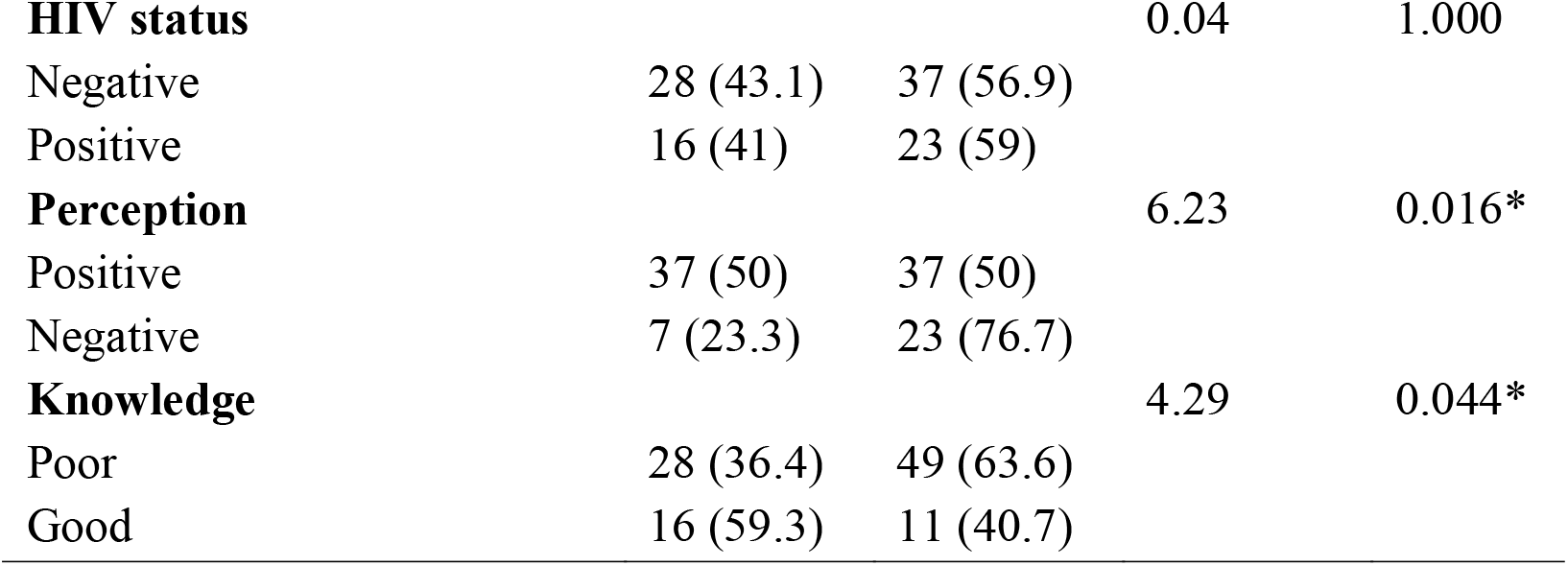
Association between participants’ socio-demographics and hesitancy to COVID-19 vaccine.

### Predictors of COVID-19 Vaccine Hesitancy among Participants

Table 3 presents the factors predicting COVID-19 vaccine hesitancy among participants. Participants who are parous (aOR=3.31, p=0.024, CI: 1.17-9.42) were more likely to be hesitant to COVID-19 vaccine compared to those who are nulliparous. Healthy participants (aOR=3.95, p=0.003, CI: 1.55-10.07) were 4 times more likely to be hesitant to COVID-19 vaccine than those who were not healthy. Participants that have family and friends that have taken the COVID-19 vaccine (aOR=3.27, p=0.037, CI: 1.07-10.00) were 3 times more likely to be hesitant compared to participants’ which have family and friends that has not taken the COVID-19 vaccine. Participants with a negative perception (aOR=3.47, p=0.004, CI: 1.16-10.34) of the COVID-19 vaccine were more likely to be hesitant to COVID-19 vaccine than those with a positive perception.

**Table 3:** Predicting factors of COVID-19 vaccine hesitancy among participants.

| Variables | AOR | 95%CI | P-value |
| --- | --- | --- | --- |
| **Parental status** |  |  |  |
| Nulliparous (ref) | 1 |  |  |
| Parous | 3.31 | 1.17-9.42 | 0.024* |
| **Travelled out the Residential area during COVID-19** |  |  |  |
| Yes (ref) | 1 |  |  |
| No | 0.55 | 0.95-3.27 | 0.518 |
| **Current Health status** |  |  |  |
| Not Healthy (ref) | 1 |  |  |
| Healthy | 3.95 | 1.55-10.07 | 0.003* |
| **Family or friend that has taken the COVID-19 vaccine** |  |  |  |
| No (ref) | 1 |  |  |
| Yes | 3.27 | 1.07-10.00 | 0.037* |
| **Perception** |  |  |  |
| Positive (ref) | 1 |  |  |
| Negative | 3.47 | 1.16-10.34 | 0.004* |
| **Knowledge** |  |  |  |
| Good (ref) | 1 |  |  |
| Poor | 2.59 | 0.92-7.30 | 0.072 |

\*statistically significant, ref- reference

## Discussion

Our study assessed COVID-19 vaccine hesitancy among pregnant women attending antenatal clinic in Jos Nigeria. We found that participants have limited and poor knowledge of the COVID-19 as well as the vaccine, and half of the participants had COVID-19 vaccine hesitancy. The significant factors influencing COVID-19 vaccine hesitancy among this population were having more than one child, being healthy, and having a family member or friends who took the vaccine.

COVID-19 as with other viral diseases during pregnancy could be very severe and increase the risk of a variety of adverse birth outcomes such as preterm birth, still birth, neonatal and infant mortality and the also the risk of the virus passing to the foetus during pregnancy (Lemma et al., 2024; Wei et al., 2021). There is also associated maternal morbidity and mortality especially among women with other co-morbidities such as diabetes, hypertension, respiratory infections among others (Gulersen et al., 2022; Villar et al., 2021). Despite these challenges of COVID-19 infection, vaccine hesitancy among pregnant women is a serious public health issue in low and middle-income countries (LMICs).

This study showed that about three-quarter of the participants got information about COVID-19 infection and prevention from the media such as Television and radio. Less than half got information from the health facilities. There is a high possibility of getting misinformation from media as opposed to health facilities because of the fake news and dearth of information that filled the media since the time of the pandemic (Gerbina, 2021). Health care providers are central to providing pregnant women with necessary and adequate information about COVID-19 infection and prevention especially during antenatal visits. This is critical to reducing infection and its complications such as adverse pregnancy and neonatal outcomes such as preterm and neonatal death and poor maternal outcomes (Lemma et al., 2024; Villar et al., 2021).

Furthermore, our study showed that the awareness and knowledge of symptoms of COVID-19 infection among the participants were varied, as the majority of the participants were aware of cough and fever as symptoms, and less than half reported difficulty with breathing, fatigue, and sore throat as symptoms of COVID-19. This is similar to a study on knowledge, attitude and practices of pregnant women related to COVID□19 infection in seven countries by (Naqvi et al., 2022). However, about three-quarter of the participants in our study reported poor knowledge of COVID-19 vaccine. With the poor knowledge of the vaccine, there is a high likelihood that pregnant women will not be willing to get the vaccine. Globally, prenatal vaccination is a proven and low-cost preventive measure that has helped to reduce the burden of infectious disease (Fleming-Dutra et al., 2023; Wu et al., 2023), such as the successful roll out of tetanus toxoid in Nigeria. The COVID-19 vaccine is recommended for pregnant women, breastfeeding women, or any woman who plans to get pregnant (Kalafat et al., 2022). It is a critical strategy to control the spread and reduce the severity of COVID-19 infection (Bhattacharya et al., 2022). However, pregnant women’s limited knowledge of COVID-19 and vaccination can increase the risk of associated complications (Naqvi et al., 2022). It is therefore essential for pregnant women to have accurate information about COVID-19 for a robust preventive measure.

About half of the participants were hesitant to take up the COVID-19 vaccines in this study. According to the World Health Organization, vaccine hesitancy is defined as ‘a delay in accepting or refusing immunisation notwithstanding the availability of vaccination services (World Health Organization, 2014). In a cross-sectional study by Comparcini et al to assess the factors influencing COVID-19 vaccine hesitancy among pregnant and breastfeeding/puerperium women in Italy, a high vaccine hesitancy rate was reported among the participants (Comparcini et al., 2024). Our study showed that significantly, the likelihood of COVID-19 vaccine hesitancy reported among participants was having one or more children, being in a healthy state, having family and friends who have taken the COVID-19 vaccine, and having a negative perception. Several studies have reported varying factors influencing vaccine hesitancy among pregnant women, especially in LIMC such as Nigeria. The frequently reported reasons contributing to vaccine hesitancy in other studies were misinformation about the vaccine’s safety for the mother and unborn child, religious/cultural beliefs, lack of trust/confidence in government and healthcare providers (Adeyanju et al., 2022; Ogbuabor & Chime, 2021; Omunakwe et al., 2024).

However, several studies have documented the safety of COVID-19 vaccination during pregnancy, and data support protection extending to infants through maternal antibody transfer (Villar et al., 2023; Yussuph et al., 2023). Evidence from systematic reviews and meta-analysis showed that COVID-19 vaccine significantly lowered risk of preterm birth, and severe illness during pregnancy (Prasad et al., 2022). A case control study by Schrag et al in a study in the USA reported effectiveness of COVID-19 vaccine in reducing hospitalization as a result of COVID-19 infection among vaccinated compared to unvaccinated pregnant women with COVID-19 infection (Schrag et al., 2022). Several cohort studies have reported that COVID-19 vaccine reduced the occurrence of stillbirth and preterm birth, with no adverse impact on fetal growth or development (Hui et al., 2023; Villar et al., 2023).

### Study limitation

This study is limited in a number of ways. First, it was limited to pregnant women receiving prenatal care at a state hospital in Jos, which may limit the findings’ applicability to other parts of Nigeria with distinct sociodemographic and cultural contexts. Second, social desirability and recall biases may have affected self-reported data, which could affect how accurately replies on vaccine views and past infections are obtained.

## Conclusion

Our study highlights a high COVID-19 vaccine hesitancy among pregnant women in Jos, Nigeria, driven largely by poor knowledge of the disease and the vaccine, misinformation, and personal perceptions of health risk. Despite evidence supporting the safety and effectiveness of COVID-19 vaccination in pregnancy, hesitancy remains a major barrier to prenatal vaccination. Strengthening antenatal health education, improving communication between healthcare providers and pregnant women, and addressing misinformation through trusted channels are critical to increasing vaccine acceptance and uptake. One reason for higher vaccine hesitancy in pregnant women, particularly to COVD-19, apart from having a family member report back some vaccine related adverse events, may be attributed to late roll out of these vaccines of pregnant women. Pregnant women are usually excluded from clinical or vaccine trials which further impacts vaccine updates and may lead to higher vaccine hesitancy in this population. Integrating targeted health promotion strategies into existing maternal health programs can help reduce vaccine hesitancy, improve vaccine uptake, and ultimately protect both mothers and their infants from vaccine preventable diseases.

## Data Availability

All data produced in the present study are available upon reasonable request to the authors

## Notes

### Competing Interest Statement

The authors have declared no competing interest.

### Funding Statement

This study was funded by NIH 3U01HD094658-05S1

### Author Declarations

Plateau state Specialist Hospital Jos Local institutational review board.

## References

Adeyanju, G. C., Sprengholz, P., & Betsch, C. (2022). Understanding drivers of vaccine hesitancy among pregnant women in Nigeria: A longitudinal study. NPJ Vaccines, 7(1), 96. 10.1038/s41541-022-00489-7

Alirezaylavasani, A., Skeie, L. G., Egner, I. M., Chopra, A., Dahl, T. B., Prebensen, C., Vaage, J. T., Halvorsen, B., Lund-Johansen, F., Tonby, K., Reikvam, D. H., Stiksrud, B., Holter, J. C., Dyrhol-Riise, A. M., Munthe, L. A., & Kared, H. (2024). Vaccine responses and hybrid immunity in people living with HIV after SARS-CoV-2 breakthrough infections. Npj Vaccines, 9(1), 1–15. 10.1038/s41541-024-00972-3

Ashkir, S., Abel, T., Khaliq, O. P., & Moodley, J. (2023). COVID-19 vaccine hesitancy among pregnant women in an antenatal clinic in Durban, South Africa. Southern African Journal of Infectious Diseases, 38(1), a516. 10.4102/sajid.v38i1.516

Basoulis, D., Mastrogianni, E., Voutsinas, P. M., & Psichogiou, M. (2023). HIV and COVID-19 Co-Infection: Epidemiology, Clinical Characteristics, and Treatment. Viruses, 15(2), 577. 10.3390/V15020577

Bhattacharya, O., Siddiquea, B. N., Shetty, A., Afroz, A., & Billah, B. (2022). COVID-19 vaccine hesitancy among pregnant women: a systematic review and meta-analysis. BMJ Open, 12(8), e061477. 10.1136/bmjopen-2022-061477

Comparcini, D., Tomietto, M., Pastore, F., Nichol, B., Miniscalco, D., Flacco, M. E., Stefanizzi, P., Tafuri, S., Cicolini, G., & Simonetti, V. (2024). Factors Influencing COVID-19 Vaccine Hesitancy in Pregnant and Breastfeeding/Puerperium Women: A Cross-Sectional Study. Vaccines, 12, 772. 10.3390/vaccines12070772

Dagan, N., Barda, N., Biron-Shental, T., Makov-Assif, M., Key, C., Kohane, I. S., Hernán, M. A., Lipsitch, M., Hernandez-Diaz, S., Reis, B. Y., & Balicer, R. D. (2021). Effectiveness of the BNT162b2 mRNA COVID-19 vaccine in pregnancy. Nature Medicine, 27(10), 1693–1695. 10.1038/s41591-021-01490-8

Dirie, N. I., Abdullahi, M., Nur, S., Mohamud, A. K., Garba, B., Dahie, H. A., Adam, M. H., & Mohamoud, J. H. (2024). COVID-19 Vaccine Uptake and Factors Associated Among Pregnant Women in Mogadishu, Somalia. Infection and Drug Resistance, 17, 3933–3943. 10.2147/IDR.S471674

Fleming-Dutra, K. E., Zauche, L. H., Roper, L. E., Ellington, S. R., Olson, C. K., Sharma, A. J., Woodworth, K. R., Tepper, N., Havers, F., Oliver, S. E., Twentyman, E., & Jatlaoui, T. C. (2023). Safety and Effectiveness of Maternal COVID-19 Vaccines Among Pregnant People and Infants. Obstetrics and Gynecology Clinics of North America, 50(2), 279–297. 10.1016/J.OGC.2023.02.003

Gerbina, T. V. (2021). Science Disinformation: On the Problem of Fake News. In Scientific and Technical Information Processing (Vol. 48, Issue 4, pp. 290–298). 10.3103/S0147688221040092

Ghamri, R. A., Othman, S. S., Alhiniah, M. H., Alelyani, R. H., Badawi, A. M., & Alshahrani, A. A. (2022). Acceptance of COVID-19 Vaccine and Associated Factors Among Pregnant Women in Saudi Arabia. Patient Preference and Adherence, 16, 861–873. 10.2147/PPA.S357653

Gulersen, M., Rochelson, B., Shan, W., Wetcher, C. S., Nimaroff, M., & Blitz, M. J. (2022). Severe maternal morbidity in pregnant patients with SARS-CoV-2 infection. American Journal of Obstetrics & Gynecology MFM, 4(4), 100636. 10.1016/j.ajogmf.2022.100636

Haynes, B. F. (2021). A New Vaccine to Battle Covid-19. New England Journal of Medicine, 384(5), 470–471. 10.1056/NEJME2035557

Hui, L., Marzan, M. B., Rolnik, D. L., Potenza, S., Pritchard, N., Said, J. M., Palmer, K. R., Whitehead, C. L., Sheehan, P. M., Ford, J., Mol, B. W., & Walker, S. P. (2023). Reductions in stillbirths and preterm birth in COVID-19-vaccinated women: a multicenter cohort study of vaccination uptake and perinatal outcomes. American Journal of Obstetrics and Gynecology, 228(5), 585.e1-585.e16. 10.1016/j.ajog.2022.10.040

Kalafat, E., Heath, P., Prasad, S., OBrien, P., & Khalil, A. (2022). COVID-19 vaccination in pregnancy. American Journal of Obstetrics and Gynecology, 227(2), 136. 10.1016/J.AJOG.2022.05.020

Lemma, T., Silesh, M., Taye, B. T., Desta, K., Moltot, T., Melisew, A., Sisay, M., Zeneb, W., & Dagnaw, Y. (2024). Knowledge, attitude and practice towards COVID-19 among pregnant women in Africa: A systematic review and meta-analysis. Heliyon, 10(11), e31926. 10.1016/j.heliyon.2024.e31926

MacDonald, N. E., & the SAGE Working Group on Vaccine Hesitancy. (2015). Vaccine hesitancy: Definition, scope and determinants. Vaccine, 33(34), 4161–4164. 10.1016/j.vaccine.2015.04.036

Naqvi, F., Naqvi, S., Billah, S. M., Saleem, S., Fogleman, E., Peres-da-Silva, N., Figueroa, L., Mazariegos, M., Garces, A. L., Patel, A., Das, P., Kavi, A., Goudar, S. S., Esamai, F., Chomba, E., Lokangaka, A., Tshefu, A., Haque, R., Siraj, S., … Goldenberg, R. L. (2022). Knowledge, attitude and practices of pregnant women related to COVID-19 infection: A cross-sectional survey in seven countries from the Global Network for Women’s and Children’s Health. BJOG___J: An International Journal of Obstetrics and Gynaecology, 129(8), 1289–1297. 10.1111/1471-0528.17122

Nomah, D. K., Reyes-Urueña, J., Llibre, J. M., Ambrosioni, J., Ganem, F. S., Miró, J. M., & Casabona, J. (2022). HIV and SARS-CoV-2 Co-infection: Epidemiological, Clinical Features, and Future Implications for Clinical Care and Public Health for People Living with HIV (PLWH) and HIV Most-at-Risk Groups. Current HIV/AIDS Reports, 19(1), 17–25. 10.1007/S11904-021-00596-5

Ogbuabor, D. C., & Chime, A. C. (2021). Prevalence and predictors of vaccine hesitancy among expectant mothers in Enugu metropolis, South-east Nigeria. Journal of Public Health Policy, 42(2), 222–235. 10.1057/s41271-020-00273-8

Omunakwe, H. E., Okuku, M., Amadi, S. C., & Dan-Jumbo, A. (2024). COVID-19 vaccine acceptance among pregnant women: a multicentre cross-sectional survey in Port Harcourt, Nigeria. The Pan African Medical Journal, 47, 72. 10.11604/pamj.2024.47.72.37446

Prasad, S., Kalafat, E., Blakeway, H., Townsend, R., O’Brien, P., Morris, E., Draycott, T., Thangaratinam, S., Le Doare, K., Ladhani, S., von Dadelszen, P., Magee, L. A., Heath, P., & Khalil, A. (2022). Systematic review and meta-analysis of the effectiveness and perinatal outcomes of COVID-19 vaccination in pregnancy. Nature Communications, 13(1), 2414. 10.1038/s41467-022-30052-w

Schrag, S. J., Verani, J. R., Dixon, B. E., Page, J. M., Butterfield, K. A., Gaglani, M., Vazquez-Benitez, G., Zerbo, O., Natarajan, K., Ong, T. C., Lazariu, V., Rao, S., Beaver, R., Ellington, S. R., Klein, N. P., Irving, S. A., Grannis, S. J., Kiduko, S., Barron, M. A., … Naleway, A. L. (2022). Estimation of COVID-19 mRNA Vaccine Effectiveness Against Medically Attended COVID-19 in Pregnancy During Periods of Delta and Omicron Variant Predominance in the United States. JAMA Network Open, 5(9), e2233273–e2233273. 10.1001/jamanetworkopen.2022.33273

Toubasi, A. A., Al-Sayegh, T. N., Obaid, Y. Y., Al-Harasis, S. M., & AlRyalat, S. A. S. (2022). Efficacy and safety of COVID□19 vaccines: A network meta□analysis. Journal of Evidence-Based Medicine, 15(3), 245. 10.1111/JEBM.12492

Villar, J., Ariff, S., Gunier, R. B., Thiruvengadam, R., Rauch, S., Kholin, A., Roggero, P., Prefumo, F., do Vale, M. S., Cardona-Perez, J. A., Maiz, N., Cetin, I., Savasi, V., Deruelle, P., Easter, S. R., Sichitiu, J., Soto Conti, C. P., Ernawati, E., Mhatre, M., … Papageorghiou, A. T. (2021). Maternal and Neonatal Morbidity and Mortality Among Pregnant Women With and Without COVID-19 Infection: The INTERCOVID Multinational Cohort Study. JAMA Pediatrics, 175(8), 817–826. 10.1001/jamapediatrics.2021.1050

Villar, J., Soto Conti, C. P., Gunier, R. B., Ariff, S., Craik, R., Cavoretto, P. I., Rauch, S., Gandino, S., Nieto, R., Winsey, A., Menis, C., Rodriguez, G. B., Savasi, V., Tug, N., Deantoni, S., Fabre, M., Martinez de Tejada, B., Rodriguez-Sibaja, M. J., Livio, S., … Papageorghiou, A. T. (2023). Pregnancy outcomes and vaccine effectiveness during the period of omicron as the variant of concern, INTERCOVID-2022: a multinational, observational study. Lancet (London, England), 401(10375), 447–457. 10.1016/S0140-6736(22)02467-9

Wei, S. Q., Bilodeau-Bertrand, M., Liu, S., & Auger, N. (2021). The impact of COVID-19 on pregnancy outcomes: a systematic review and meta-analysis. CMAJ, 193(16), E540–E548. 10.1503/cmaj.202604

World Health Organization. (2014). The Strategic Advisory Group of Experts (SAGE). Report of the SAGE Working group on Vaccine Hesitancy.

World Health Organization. (2025a). COVID-19 deaths. WHO COVID-19 Dashboard.

World Health Organization. (2025b). Vaccines and immunization.

Wu, S., Wang, L., Dong, J., Bao, Y., Liu, X., Li, Y., Liu, X., Xie, H., & Ying, H. (2023). The dose- and time-dependent effectiveness and safety associated with COVID-19 vaccination during pregnancy: a systematic review and meta-analysis. International Journal of Infectious Diseases, 128, 335–346. 10.1016/j.ijid.2023.01.018

Yussuph, Z. H., Alwy Al-Beity, F. M., August, F., & Anaeli, A. (2023). COVID-19 vaccine hesitancy among pregnant women attending public antenatal clinics in Dar es Salaam, Tanzania. Human Vaccines and Immunotherapeutics, 19(3), 2269777. 10.1080/21645515.2023.2269777

